# Association between Stevens-Johnson syndrome and toxic epidermal necrolysis with ibuprofen: A pharmacovigilance study in the UK Yellow Card scheme and systematic review of case reports

**DOI:** 10.1101/2023.12.05.23299283

**Authors:** Guy Fletcher, David K. Ryan, C B. Bunker

**Author notes:** **Corresponding author:** C B Bunker.

## Abstract

**Introduction:** Stevens-Johnson syndrome (SJS) and toxic epidermal necrolysis (TEN) are a group of severe acute muco-cutaneous blistering disorders with a significant burden of morbidity and mortality. Drugs are commonly identified as potential precipitants of SJS/TEN, although it can be difficult to firmly identify causative agents. Ibuprofen has been proposed as a rare trigger for SJS/TEN and given the widespread use of this non-steroidal anti-inflammatory and significance of reaction, further pharmacovigilance analysis is warranted.

**Methods:** Serious or fatal adverse drug reaction (ADR) data from 1998 – October 2023 were downloaded from the UK MHRA Yellow Card Scheme website. Proportional reporting ratios (PRR) and 95% confidence intervals were calculated for SJS and TEN associated with ibuprofen using standard pharmacovigilance methods. Secondly, a systematic review of literature was conducted to explore reported cases where ibuprofen was the suspected trigger for a case of SJS/TEN. Eligible cases were assessed for the likelihood of ibuprofen being the causative agent by two reviewers using the Naranjo adverse drug reaction probability scale and the ALDEN algorithm for the assessment of drug causality in SJS/TEN.

**Results:** In total, 1, 994, 967 ARDs, relating to 1,811 SJS reports and 1,299 TEN reports. There was an increased signal for reports of SJS-TEN associated with ibuprofen use. The PRR for SJS with ibuprofen was 3.73 (95% confidence interval 2.68 – 5.18) and 4.49 (95% confidence interval 3.15 – 6.41). In our literature review, 23 individual case reports were deemed eligible for inclusion and the majority of these cases were determined as being doubtful in causality assessment. Difficulty in attributing causality was related to poorly defined temporal relationships between ibuprofen and SJS-TEN or poor reporting standards of case reports.

**Conclusions:** There are pharmacovigilance signals suggesting increased reporting of SJS/TEN in patients exposed to ibuprofen within UK data. However, more detailed case reports describing this association in literature have low likelihood of supporting causality. It is possible that reverse causality or co-causality is mediating this association. Further studies and pharmacovigilance is required to clarify the association between SJS/TEN and ibuprofen.

## Introduction

Stevens-Johnson syndrome (SJS) and toxic epidermal necrolysis (TEN) are part of a spectrum of severe acute muco-cutaneous blistering disorders with a significant burden of morbidity and mortality. Drugs are commonly identified as potential precipitants of SJS-TEN, although it can be difficult to ascertain causative agents. Ibuprofen has been proposed as a trigger for SJS/TEN. Given the widespread use of ibuprofen, further pharmacovigilance analysis is warranted.

Our aims were two-fold, firstly to explore pharmacovigilance signals for association of ibuprofen with SJS-TEN in the UK national adverse drug reporting Yellow Card scheme. Secondly, we aimed to appraise case reports of SJS-TEN with ibuprofen for likelihood of this agent being causal for an adverse drug reaction (ADR) using standardised criteria. In this way, we sought to further explore the relationship between SJS/TEN and ibuprofen.

## Methods

Serious or fatal ADRs from 1998 – October 2023 were obtained from the UK Yellow Card Scheme. Proportional reporting ratios (PRR) and 95% confidence intervals were calculated for SJS and TEN associated with ibuprofen (supplementary methods 1). A PRR > 1 implies that the signal for SJS/TEN is more commonly reported for ibuprofen, compared with background reporting signals.

Secondly, a systematic review of literature was conducted to explore reported cases where ibuprofen was the suspected trigger for a case of SJS/TEN. Medline and EMBASE data bases were accessed on 31/10/23 for English language case series or reports (supplementary methods 2). Eligible cases were assessed for the likelihood of ibuprofen being the causative agent using the Naranjo adverse drug reaction probability scale [2] and the ALDEN algorithm [3] for the assessment of drug causality in SJS/TEN (supplementary methods 3). These frameworks provide two different standardised approaches for assessing the likelihood that a specific drug might be the causal agent in producing an ADR.

## Results

In total, 1, 994, 967 ARDs were analysed, comprising 1, 811 SJS reports and 1, 299 TEN reports. Out of the 10,802 reports relating to ibuprofen, there were 36 cases of SJS and 31 cases of TEN. The PRR showed disproportionately high signals for SJS and TEN with ibuprofen compared to other drugs. The PRR for SJS with ibuprofen was 3.73 (95% confidence interval 2.68 – 5.18) and 4.49 (95% confidence interval 3.15 – 6.41).

In our literature review, 23 individual case reports were deemed eligible for inclusion. The majority (14/23) of these cases were assigned by two authors independently as being ‘doubtful’ for a causative relationship on the Naranjo scale, with the remaining being ‘possible’. Similar findings were found using the ALDEN algorithm for a causative drug agent in SJS (13/23 – ‘unlikely or very unlikely’ causal relationship, 9/19 – ‘possible’ and 1/19 – ‘probable’).

## Discussion

UK SJS-TEN guidelines [4] advocate the early recognition and swift withdrawal of the offending drug to reduce morbidity and mortality and should form part of routine care. Therefore, an understanding of the association between common drugs and SJS-TEN is essential.

Ibuprofen is one of the most utilised over-the-counter analgesic agents and has been suggested as a potential trigger for SJS-TEN. In this study, there are signals for increased reporting of association with ibuprofen and SJS-TEN in the UK national ADR database. However, this is based on signals from spontaneous reports and are limited by data availability, recall bias and confounding factors. It is not possible to infer causal relationship from this data. Secondly, a systematic literature review suggests that causality cannot be inferred from all published reports. This may reflect poor reporting standards in case reports but could also reflect the complex association between SJS-TEN and ibuprofen. Also the Naranjo and ALDEN systems do not lend themselves suitably to all clinical scenarios and all suspect drugs. For example, difficulties in attributing causality were related to defining the temporal relationship between ibuprofen and SJS-TEN. Inescapably, the total number of ‘possibles’ and ‘probable’ using the Naranjo and ALDEN scales does leave a cloud of suspicion over the drug.

It is possible that several mechanisms could be mediating such an observation (Figure 1). Reverse causality could be creating the association between ibuprofen and SJS-TEN, whereby the prodromal symptoms of SJS-TEN triggers utilisation of ibuprofen. Co-causation could also be responsible, whereby the causative agent for SJS-TEN is a viral agent and this creates an immunological milieu, that triggers or amplifies the development of an ADR to ibuprofen. This is analogous to other ADRs such as the cutaneous ADR that can complicate amoxicillin in glandular fever/Epstein-Barr virus infection and human herpes virus 6 & 7 reactivation in DRESS (drug reaction with eosinophilia and systemic symptoms).[5,6]

**Figure 1:**
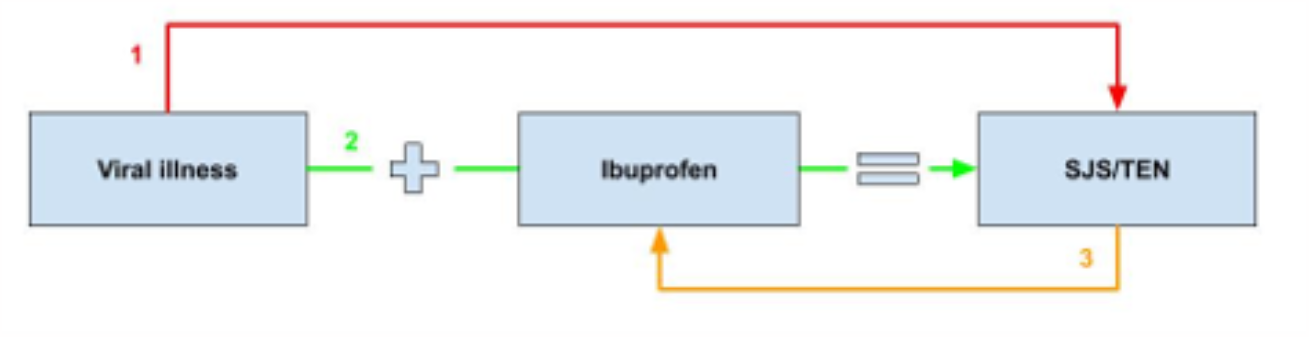
Directed acyclic graph showing potential association between SJS/TEN, ibuprofen and viral illnesses. Figure 1: Directed acyclic graph showing possible associations between the observed pharmacovigilance signal suggesting increased reports of SJS-TEN with ibuprofen use. 1. **Confounding**: Ibuprofen may be a confounding factor, that relates to the exposure (viral illness or other precipitant) and the outcome, SJS but not on the causal pathway. | 2. **Co-causal relationship**: Both a viral illness and ibuprofen are required to trigger SJS-TEN. 3. **Reverse causality:** The prodrome of SJS-TEN results in an individual taking ibuprofen for symptomatic relief, with no causal relationship between ibuprofen and SJS-TEN.

SJS-TEN is known to begin with prodromal symptoms, that are difficult to differentiate from other possible diagnostic considerations, including viral prodromes. As ibuprofen may be used for symptomatic relief of the prodromal-phase of SJS-TEN, ibuprofen may not necessarily reflect the cause of SJS-TEN. Caution must therefore be taken in attributing causality in this context, but our work tends to incriminate rather than exculpate ibuprofen in SJS/TEN.

## Conclusion

There are pharmacovigilance signals suggesting increased reporting of SJS-TEN in patients exposed to ibuprofen. Furthermore, a systematic review has suggested a complex relationship between ibuprofen and SJS-TEN. Possibilities are that reverse causality is mediating this association, infection may be a confounder, or an infection and ibuprofen are mutually dependent causative co-factors in SJS/TEN.

## Data Availability

All data produced are available online at the Yellow Card Scheme, MHRA, UK.

https://yellowcard.mhra.gov.uk/

## Supplementary methods 1

The Medicines and Healthcare Products Regulatory Agency collect adverse drug reaction (ADR) for the UK via the Yellow Card Scheme. This database was accessed on the 18^th^ October 2023 to download all serious and fatal reports relating to a drug. Disproportionality analysis was conducted on this database to determine which drugs have higher signals for an index symptom or event, compared to background signals. The formula and 95% confidence intervals for the proportional risk ratio were calculated using standardised pharmacovigilance methods:

- The value A indicates the number of individual cases with the suspect medicinal product P involving an adverse event R.
- The value 13 indicates the number of individual cases related to the suspect medicinal product P, involving any other adverse events but R.
- The value C indicates the number of individual cases involving event R in relation to any other medicinal products but P.
- The value D indicates the number of individual cases involving any other adverse events but R and any other medicinal products but P.

The proportional reporting ratio (PRR) for each adverse event R:

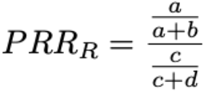

The standard deviation S for the PRR is calculated using the formula:

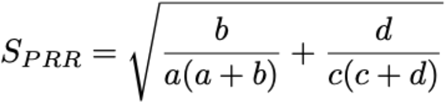

The corresponding 95% confidence interval is derived using:

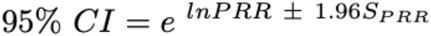

## Supplementary methods 2

A systematic review of case studies, case series and systematic reviews relating to SJS-TEN and ibuprofen was conducted on 31/10/23. The following databases were searched: Medline and EMBASE

The search strategy for Medline was:

| # ▲ Searches | Results |
| --- | --- |
| 1 ibuprofen/ | 10111 |
| 2 ibuprofen.mp. [mp=title, book title, abstract, original title, name of substance word, subject heading word, floating sub-heading word, keyword heading word, organism supplementary concept word, protocol supplementary concept word, rare disease supplementary concept word, unique identifier, synonyms, population supplementary concept word, anatomy supplementary concept word] | 17769 |
| 3 1 or 2 | 17769 |
| 4 toxic epidermal necrolysis.mp. [mp=title, book title, abstract, original title, name of substance word, subject heading word, floating sub-heading word, keyword heading word, organism supplementary concept word, protocol supplementary concept word, rare disease supplementary concept word, unique identifier, synonyms, population supplementary concept word, anatomy supplementary concept word] | 4293 |
| 5 exp toxic epidermal necrolysis/ | 6057 |
| 6 Steven's Johnson Syndrome.mp. [mp=title, book title, abstract, original title, name of substance word, subject heading word, floating sub-heading word, keyword heading word, organism supplementary concept word, protocol supplementary concept word, rare disease supplementary concept word, unique identifier, synonyms, population supplementary concept word, anatomy supplementary concept word] | 7633 |
| 7 Stevens Johnson syndrome/ | 6057 |
| 8 4 or 5 or 6 or 7 | 8146 |
| 9 3 and 8 | 51 |
| 10 case.mp. [mp=title, book title, abstract, original title, name of substance word, subject heading word, floating sub-heading word, keyword heading word, organism supplementary concept word, protocol supplementary concept word, rare disease supplementary concept word, unique identifier, synonyms, population supplementary concept word, anatomy supplementary concept word] | 3945484 |
| 11 9 and 10 | 39 |

A similar search strategy was used for EMBASE and produced 81 results.

### Exclusion criteria

Cases were excluded if they were not relevant, not in English language or they failed to provide enough evidence to meaningfully answer ADLEN or Naranjo criteria. Duplicates between the two databases were used once.

Abstracts were reviewed manually by two authors to ensure relevance.

97 articles were excluded based on the criteria above, resulting in 18 articles with 23 individual cases.

## Supplementary methods 3

All case studies relating to ibuprofen and SJS-TEN were reviewed independently by two authors (GF and DKR) according to the Narajo and ALDEN criteria. Any disagreement was resolved through discussion between the two assessors. Both assessors are medically trained and assessor, GF has experience in Dermatology with the other author, DKR being experienced in Clinical Pharmacology.

The Narajo criteria is a standardised causality assessment tool for all adverse drug reactions. It was initially intended for use in controlled medication trials but has found use in routine clinical practice due to its simplicity.

The ALDEN criteria is specifically used to assess for causality in cases of SJS-TEN.

**Table 1.** Demonstrating the Naranjo Score Calculation Method.

| **Naranjo Score** |  |  |  |
| --- | --- | --- | --- |
| **Question** | **Yes** | **No** | **Do Not Know or Not Done** |
| Are there previous conclusive repots on this reaction? | +1 | 0 | 0 |
| Did the adverse event appear after the suspected drug was given? | +2 | 0 | 0 |
| Did the adverse reaction improved when the drug was discontinued, or a specific antagonist was given? | +1 | 0 | 0 |
| Did the adverse reaction appear when the drug was readministered? | +2 | -1 | 0 |
| Are there alternative causes that could have caused the reaction? | -1 | +2 | 0 |
| Did the reaction reappear when a placebo was given? | -1 | +1 | 0 |
| Was the drug detected in any body fluid in toxic concentrations? | +1 | 0 | 0 |
| Was the reaction more severe when the dose was increased, or less severe when the dose was decreased? | +1 | 0 | 0 |
| Did the patient have a similar reaction to the same or similar drugs in any previous exposure? | +1 | 0 | 0 |
| Was the adverse event confirmed by objective evidence? | +1 | 0 | 0 |
| </=0 doubtful, 1-4 possible, 5-8 probable, >= 9 definite |  |  |  |

**Table 2.**
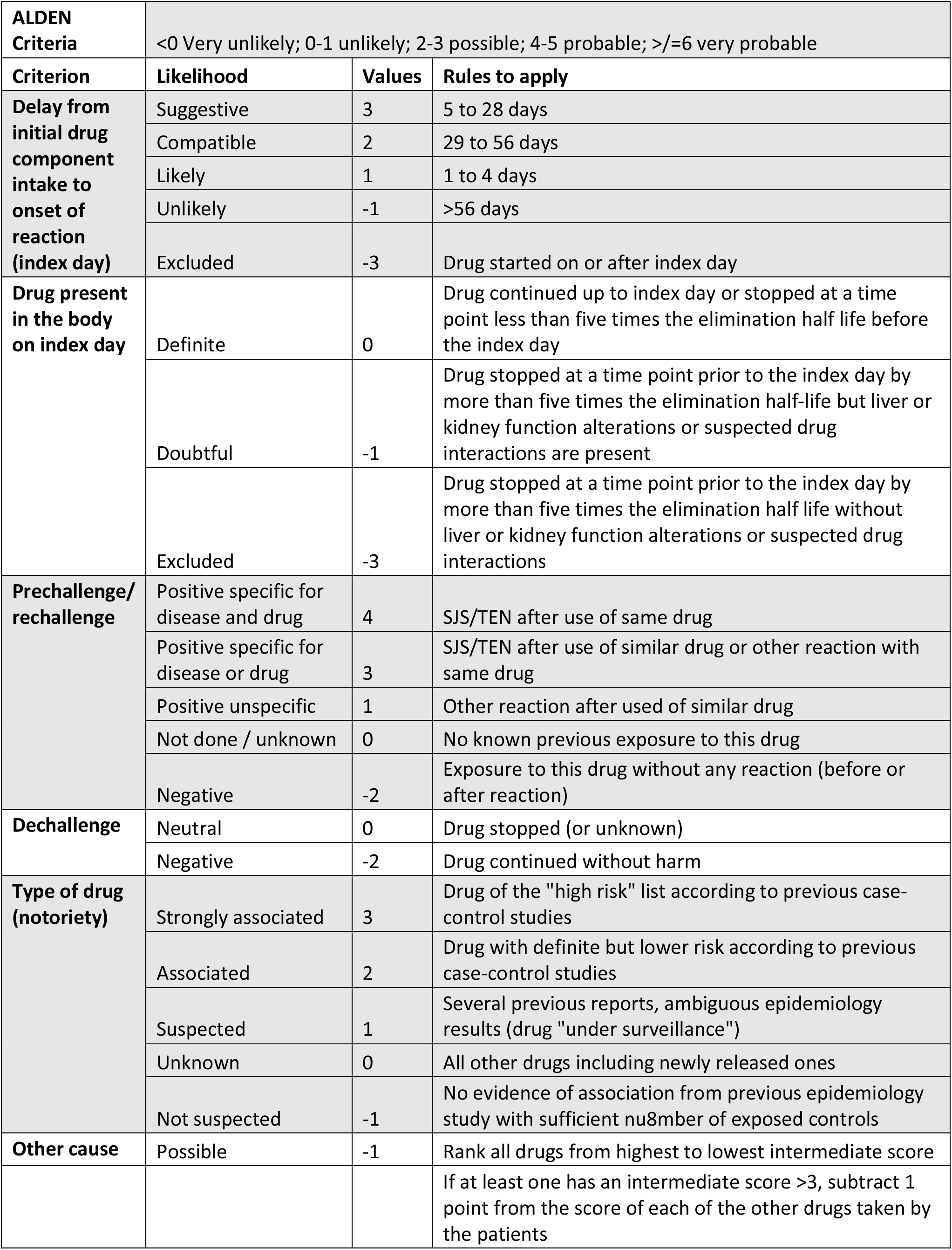
Demonstrating the ALDEN Score Calculation Method.

| **ALDEN Criteria** | <0 Very unlikely; 0-1 unlikely; 2-3 possible; 4-5 probable; >=6 very probable |  |  |
| --- | --- | --- | --- |
| **Criterion** | **Likelihood** | **Values** | **Rules to apply** |
| **Delay from initial drug component intake to onset of reaction (index day)** | Suggestive | 3 | 5 to 28 days |
|  | Compatible | 2 | 29 to 56 days |
|  | Likely | 1 | 1 to 4 days |
|  | Unlikely | -1 | >56 days |
|  | Excluded | -3 | Drug started on or after index day |
| **Drug present in the body on index day** | Definite | 0 | Drug continued up to index day or stopped at a time point less than five times the elimination half life before the index day |
|  | Doubtful | -1 | Drug stopped at a time point prior to the index day by more than five times the elimination half-life but liver or kidney function alterations or suspected drug interactions are present |
|  | Excluded | -3 | Drug stopped at a time point prior to the index day by more than five times the elimination half life without liver or kidney function alterations or suspected drug interactions |
| **Prechallenge/rechallenge** | Positive specific for disease and drug | 4 | SJS/TEN after use of same drug |
|  | Positive specific for disease or drug | 3 | SJS/TEN after use of similar drug or other reaction with same drug |
|  | Positive unspecific | 1 | Other reaction after used of similar drug |
|  | Not done / unknown | 0 | No known previous exposure to this drug |
|  | Negative | -2 | Exposure to this drug without any reaction (before or after reaction) |
| **Dechallenge** | Neutral | 0 | Drug stopped (or unknown) |
|  | Negative | -2 | Drug continued without harm |
| **Type of drug (notoriety)** | Strongly associated | 3 | Drug of the "high risk" list according to previous case-control studies |
|  | Associated | 2 | Drug with definite but lower risk according to previous case-control studies |
|  | Suspected | 1 | Several previous reports, ambiguous epidemiology results (drug "under surveillance") |
|  | Unknown | 0 | All other drugs including newly released ones |
|  | Not suspected | -1 | No evidence of association from previous epidemiology study with sufficient number of exposed controls |
| **Other cause** | Possible | -1 | Rank all drugs from highest to lowest intermediate score |
|  |  |  | If at least one has an intermediate score >3, subtract 1 point from the score of each of the other drugs taken by the patients |

